# Enhanced patient counselling and SMS reminder messages to improve equitable access to community-based eye care services in Meru, Kenya: An embedded, pragmatic, individual-level, two arm, equity-focused, Bayesian RCT within an adaptive platform trial

**DOI:** 10.1101/2025.04.09.25325459

**Authors:** Luke N Allen, Min Jung Kim, Michael Gichangi, David Macleod, James Carpenter, Malebogo Tlhajoane, Sarah Karanja, Nigel Bolster, Cosmas Bunywera, Hilary Rono, Francesco Merletti, Demisse Tadesse, Kennedy Odero, David Munyendo, Aphiud Njeru, Solomon Murira, Thadeus Omoga, Amos Mutinda, Matthew Burton, Andrew Bastawrous

## Abstract

**Background:** In Kenya’s Meru county, only 46% of people identified with an eye problem during screening go on to access and receive eye care services at local clinics, with younger adults (those aged 18-44) being the least likely to receive care. In previous work, our team conducted interviews with non-attenders from this ‘left-behind’ group to explore how access to essential eye services could be equitably improved. They told us that better counselling and SMS reminders would improve access.

**Methods:** We developed enhanced counselling and SMS reminders with lay input and tested this bundled intervention against standard care using a pragmatic, two-arm randomised controlled trial that was embedded within Meru’s ongoing screening programme. All consenting referred adults were enrolled. Our primary outcome was the proportion of referred 18–44-year-olds who accessed their local clinic following referral. Following our adaptive platform trial master protocol, our embedded Bayesian-based algorithm used accruing attendance data to calculate the posterior probabilities of effect difference between the arms every seven days. Participants were continually recruited until one of two stopping rules were met: there was a >95% probability that either one arm was more effective, or that the difference between the arms was <1%.

**Findings:** Our testing algorithm stopped the trial after 30 days based on analysis of outcome data from 879 people. We found a 98.6% posterior probability that the intervention arm was superior among 18-44-year-olds. The attendance rate among this group was 32.1% in the control arm vs 39.0% in the intervention arm. Secondary analyses did not show meaningful differences between the arms across the remainder of the population (i.e. adults aged >45 years).

**Discussion:** This innovative trial found evidence that an intervention bundle suggested by an underserved population group increased access to care. This embedded, adaptive, equity-focused approach has broad applications, aligned with the principle of proportionate universalism.

## Research in Context

### Evidence before this study

The WHO Thirteenth General Programme of Work states that “the main challenge to making progress towards Universal Health Coverage [UHC] comes from persistent barriers to accessing health services”. Across the African continent, approximately half of all ambulatory appointments are missed across all specialities, and sociodemographic inequalities are ubiquitous. In pursuit of UHC and the Primary Health Care principles of equity and justice, health system managers are increasingly focused on tackling unequal access to care, however the traditional approach to identifying barriers and solutions has tended to centre around expert opinion rather than engagement with affected groups. Furthermore, service modifications tend to be tested with crude ‘before-after’ methods, rather than using more robust randomised controlled trials (RCTs). In previous research, we used rapid qualitative methods to explore how young adults who had not been able to attend their clinic appointments felt services could be changed to increase access for this left-behind group. They recommended a bundled of enhanced counselling interventions.

### Added value of this study

In this study we performed a pragmatic embedded RCT, writing new code into existing community-based eye screening and referral management software so that consenting people who screened positive were automatically randomised to receive the enhanced counselling intervention or usual care at the point of referral to their local eye clinic. We used a Bayesian adaptive design, with code written into the software that examined clinic attendance data every seven days and calculated whether predetermined stopping rules had been met; based on the probability that one arm was better than the other. The trial ended after 30 days, based on a >95% probability that the intervention arm was superior. This embedded trial tested an intervention that came directly from the sociodemographic group with the lowest overall attendance rate. We found that this intervention was indeed effective, and it has now been taken to scale. The use of an adaptive design enabled the trial to run quickly; minimising the number of people allocated to the least effective arm. Automation of randomisation and statistical testing reduces the risk of human-induced bias. This approach may also reduce the barriers for implementing RCTs to test service modifications

### Implications of all the available evidence

Equitably advancing UHC is predicated on identifying and overcoming unique barriers to care, however existing efforts rarely involve consultation or co-creation with affected communities, or test the ensuing service modifications with robust methods. We have demonstrated an innovative approach for rapidly identifying which groups are being left behind; eliciting ideas for service improvements from this group; and then rigorously testing these ideas using a pragmatic, embedded, adaptive RCT design. This ‘FAIR access’ approach can be employed across a range of settings to operationalise the principles of proportionate universalism and equity-focused continuous improvement.

## Introduction

### Background and rationale

*Leave no one behind* is the ‘central promise’ of the Sustainable Development Goals^1^ and the motivating principle behind Universal Health Coverage (UHC),^2^ yet approximately one third of the world’s population still lack access to essential health services.^3^ Systematically marginalised populations often experience the worst access to essential care.^4,5^

Our research team is working to identify and address barriers to community-based essential eye services with a focus on inequalities in Botswana, India, Kenya, and Nepal.^6^ Eye care offers a microcosm for global UHC challenges: one third of the global population lacks access to basic eye care, and over a billion people are affected by visual impairment, despite the existence of highly cost-effective treatments like cataract surgery and spectacles that could restore sight for 90% of these people.^7,8^ Poor vision leads to social exclusion, poor education outcomes, reduced economic prosperity, and reduced quality of life.^7^ Damningly, evidence suggests that groups already facing social exclusion and economic hardship tend to experience the lowest rates of access to eye care – compounding socioeconomic disadvantage.^7,9^

We are using a four-stage approach to identify and address inequitable access to eye care:

1. Find people requiring care during community-based screening and document their socioeconomic characteristics;
2. Analyse attendance data to identify which groups are the least likely to receive care;
3. Interview and survey people to explore barriers and rank suggested service modifications;
4. Randomise the implementation of the most promising service modifications, as part of a data-driven, continuous improvement approach that is embedded within the local health system.

In Kenya, we have been deploying this ‘FAIR access’ approach within the ‘Vision Impact Project’ which has already reached over 1 million people using a vision testing app and referral software provided by the social enterprise Peek Vision.^10^ We have previously found that only 46% of those identified with an eye need during community-based screening in Meru County between April – July 2023, were able to access local treatment outreach clinics once referred. Age, gender, and occupation were all associated with access to care, but the greatest disparities were observed across different age groups, with only a third of younger adults (aged 18-44 years) accessing essential care compared to over half of those aged >45 years, after controlling for a range of other factors including gender, income, occupation, education, and severity of eye condition.^11^

Using rapid qualitative methods,^12^ we previously conducted 67 interviews with younger adults who were not able to access care and identified a number of barriers and ideas to improve access for this ‘left behind’ group. We then conducted a survey with representative sample of 401 additional young adults who were not able to access care and asked them to rank their peers’ ideas by likely impact. Both elements were completed within three weeks, leading to a ranked list of potential service modifications.^13^ The ranked suggestions were discussed at a workshop with representation from the programme management team, the programme funder, the programme implementing partner, the county health management team, the Ministry of Health (MoH) national office, eye charity partners, lay representatives, and the community advisory board. This group unanimously agreed that it would be feasible to implement and test a counselling intervention that bundled together five of the solutions focused on enhancing information provision at the point of referral, informing people of; (1) the treatment outreach clinic opening times, (2) the services that are available at these clinics vs those that require onward referral to hospital-based services, (3) costs involved at the outreach clinic, (4) the importance of attending, as well as (5) being sent an SMS reminder message on the day of their appointment. Further detail on the methods for package of work has been published elsewhere.^13^

The wider literature suggests that SMS reminders can play a small but important role in improving access to care.^14–16^ There is much less research on counselling and the provision of information at the point of referral. A systematic review published in JAMA in 1992 found that ‘orienting patients’ was associated with lower rates of non-attendance.^17^ More recent qualitative research with hypertensive patients in western Kenya found that inadequate counselling was a major barrier to attending clinic appointments, with lack of understanding on the importance of referrals and misconceptions about the clinics emerging as key themes.^18^ The same research team went on to develop a multi-component ‘referral adherence’ intervention that included enhanced information provision a core element.^19^ A similar ongoing study in rural Malawi is using enhanced counselling and SMS reminder messages to improve referral uptake for hearing services.^20^

In this study we aimed to test whether provision of additional information was associated with a higher probability of accessing community-based treatment outreach eye clinics in Meru, compared to standard care. We conducted this study as part of an overarching adaptive platform trial that is being used to test multiple low-risk service modifications to improve access to care, with a focus on left-behind groups.^21^

## Methods

This registered trial (ISRCTN 11329596) followed a publicly available protocol,^22^ and reported findings in line with the CONSORT guidelines for pragmatic,^23^ adaptive,^24^ parallel group,^25^ equity-focused^26^ RCTs.

### Trial Design

We used an equity-focused, Bayesian, adaptive, pragmatic, two-arm, individual-level, randomised controlled trial with 50:50 allocation, embedded within the Vision Impact Project (VIP). We embedded randomisation and allocation into the programme’s clinical management software and performed statistical testing on the accrued routinely collected referral and clinic attendance data to identify when one of two pre-defined stopping rules was reached. We focused the primary stopping rules around the group that had previously been found to have the lowest odds of accessing services; 18–44-year-olds. We refer to this group as ‘younger adults’.

### Study setting

Our trial was embedded within the VIP that is operating in seven counties in Kenya using a smartphone-based visual acuity screening app.^10,27,28^ In Meru county - the focus of our study - over 350,000 people have been screened to date, of whom 120,000 have been identified with an eye need and referred for free further assessment at local treatment outreach clinics, generally held in primary care facilities. As stated above, only a third of referred younger adults have been able to access this care. Among the general population however, the baseline rate of access is 46%.

Our trial was integrated into the screening and patient management software developed by Peek Vision, a leading provider of eye screening software that is currently being used in over 70 screening programmes across 12 low and middle income countries (LMICs).^29^ In Meru, the Peek app is being used to screen participants for vision impairment, to capture observations and sociodemographic data, and to link participants to a referral system that tracks their progression through the local eye health system. Our trial used these routinely collected data to test whether our intervention bundle was able to increase the proportion of people among those with the lowest odds of accessing services (whom we refer to as the ‘left behind’ group, in alignment with Sustainable Development Goals language) who were checked-in to triage clinics, once referred with an eye need. Embedding the trial code within the existing system obviated the need for additional data collection.

### Participants

As a pragmatic trial, the eligibility criteria were determined by the local VIP programme managers who selected which individuals would be included in the VIP screening project. We included all adults (>18 years) who participated in the eye screening programme. We excluded those who did not meet local clinical service eligibility criteria. Our original health equity analysis in Meru^30^ used data on age, gender, religion, marital status, disability, education, occupation, income, housing, assets, and health insurance to explore which characteristics were most strongly associated with access to care (sometimes called ‘referral adherence’, although we reject the lopsided emphasis this term places on individuals’ responsibility to access care rather than service providers). We included all referred adults, but focused our primary analysis on adults aged 18-44 years as this characteristic was found to be most strongly associated with poor access to care.

### Interventions

Whilst most research and service quality improvement focuses on testing interventions derived from experts and programme managers,^31^ our intervention bundle was suggested by referred patients who had not been able to access care. After our multistakeholder workshop participants had agreed on the five service modifications to test, we drafted a new verbal counselling script and SMS reminder message that included these elements. These drafts were reviewed and revised by all stakeholders. The final draft was reviewed by two further representatives from the ‘left-behind’ group. The final text was translated into Swahili and then back-translated to check that meaning had not been lost.

### Control arm: usual care referral counselling script

*“I have examined your eyes, and you have a problem, I have referred you in the system and you will receive an SMS with where and when you are supposed to attend treatment. You will come for treatment on [date] at [location], the examination will be free and you will be informed of anything else on the material day.”*

### Intervention arm: enhanced referral counselling script

*“I have found a problem with your eyes. I am referring you to the outreach treatment clinic that will be held at [location] on [date] between [time] and [time]. At the clinic, eye care professionals will perform a specialist assessment and provide any eye drops or medicines that you might need. If you need glasses, the specialists will tell you what kind you need, and what your prescription is. The assessment is completely free. Note that a small proportion of people will be found to have complex eye problems that require onward referral for hospital assessment and special lenses that cost more than standard glasses. However, the vast majority of people have their needs fully met at the outreach triage clinic and do not need hospital referral*.

*With treatment, you will be able to see more clearly. This will help with your work, seeing faces, and using your phone. It is important that you attend the clinic to protect your vision. The clinic will only be running from [day] to [day], between [hour] and [hour], so if you don’t manage to attend, you may not be able to reschedule the appointment.”*

The relevant script was read to each participant by the screener at the point of referral. The wording of the usual care counselling script was based on the screening programme training materials and observations of what screeners currently tell participants. No elements were removed i.e. this script accurately reflects usual care. Screeners do not usually read this information out to participants from a script; however, we introduced standardised wording to reduce the risk of contamination i.e. screeners delivering the same enhanced counselling elements to participants in both the intervention and control arms.

All people who were referred were sent automated SMS reminder messages on the day of referral and the day before the appointment. These messages were generated and sent by the Peek Vision platform. The content of the intervention SMS was developed by the research team in collaboration with a lay representative from the left-behind group. The messages were sent in either English or Kiswahili, depending on the participant’s preferred language.

## Control SMS Script

*Dear <<NAME>>, you were examined and found to have an eye problem. Kindly report on < <LOCATION>> on <<DATE>> for assessment. For more information contact Meru Referral Hospital.*

### Intervention SMS script

*We found that you had an eye problem. Please attend the outreach clinic at <<LOCATION>> on <<DATE>> between <<9AM-5pm>>. The specialist assessment is free If you are found to have a complex problem, you may be referred to a hospital for further care or special glasses, and this may include a fee*

*However, the vast majority of people who attend the outreach get their eye problem fixed without the need for any further referral*

*It’s important that you attend to protect your vision, and you might not have a future opportunity to access free care. See you on <<DATE>>*

The intervention SMS was sent on the day of referral, the day before referral, and on the day of the appointment. Figure 1 shows the point at which the interventions were delivered. There were no *a priori* strategies to improve adherence.

**Figure 1:**
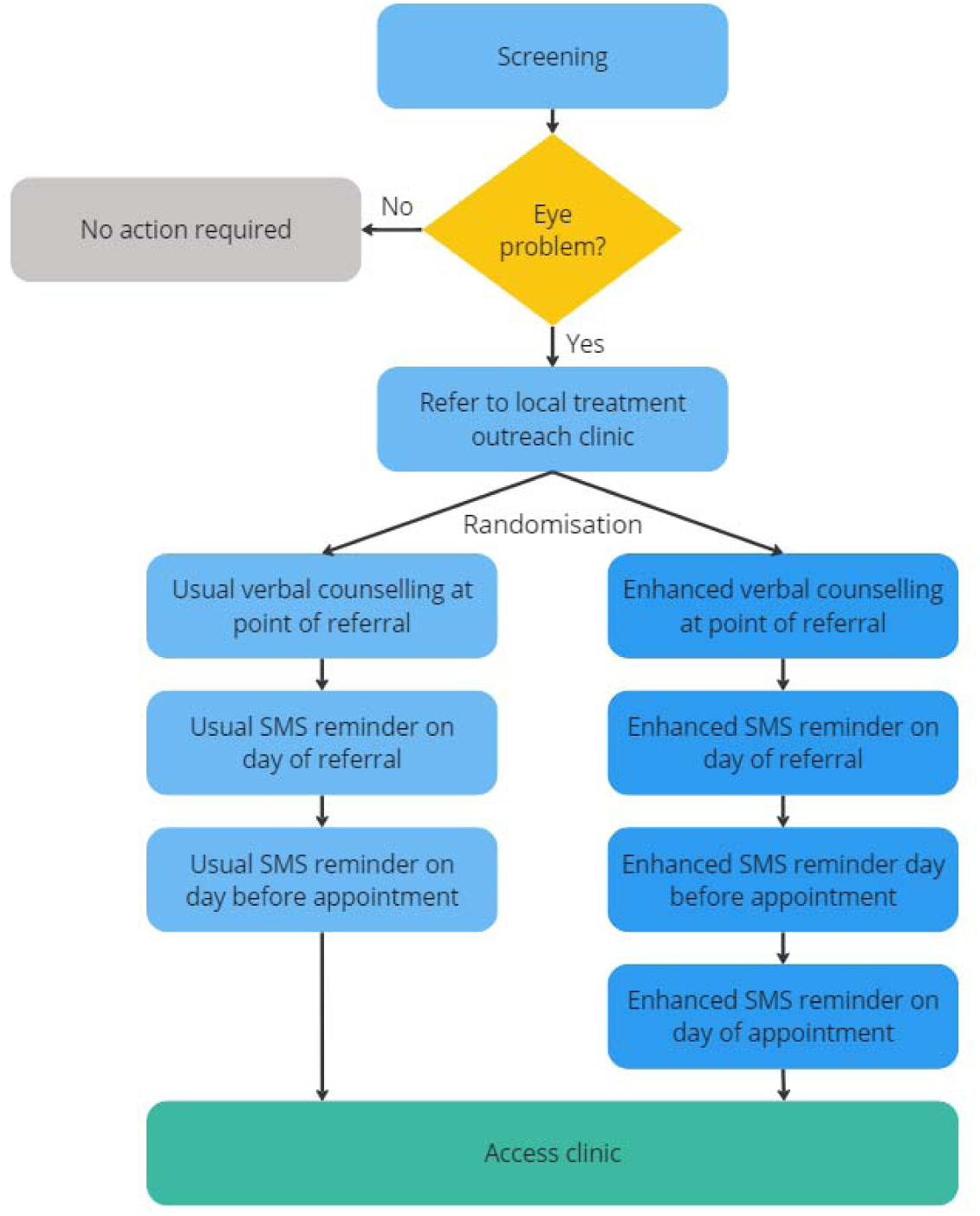
Intervention delivery

### Theory of change

Ultimately, we wanted all people with eye problems to receive the care that they need, irrespective of their sociodemographic group. In this study we assumed that those who were able to access clinics received appropriate, timely, and high-quality care. We assumed that improved vision as a result of treatment would lead to improved function, positive behavioural changes, and social and economic participation.

In previous qualitative work, people who did not manage to access care told us that an important factor that underlies the decision to seek care is the provision of adequate information around what to expect, services available, costs, time, and information about why it is important to attend. In this study, screening staff verbally provided this additional information at the point of referral. Information was also provided in follow-up SMS messages, including a message sent on the day of the appointment for those in the intervention arm. Figure 2 illustrates the causal chain, using an adaptation of the *Behaviour Centred Design* framework.^32^

**Figure 2:**
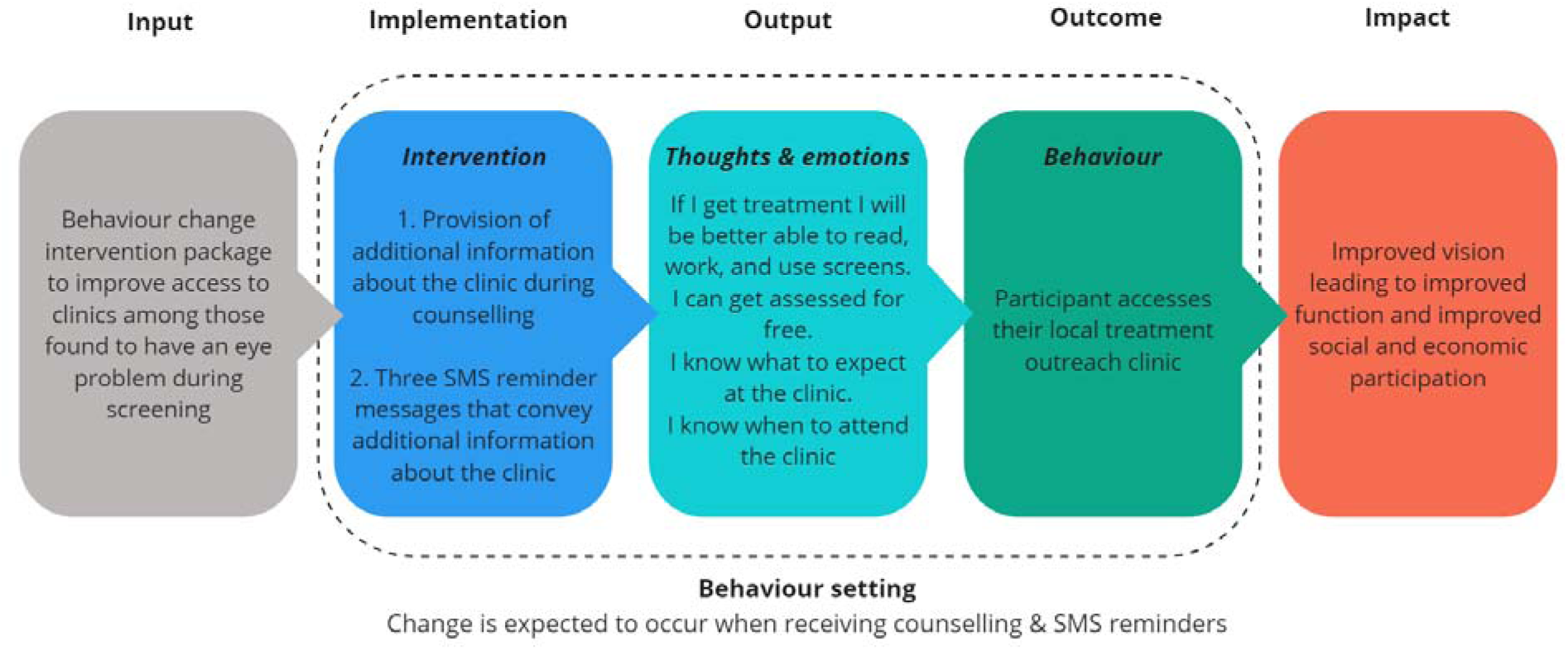
Causal pathway in our theory of change

We are the first to acknowledge that a wide range of factors influence access to care, including many critically important supply-side factors including quality, accessibility, and appropriateness.^33–35^ Furthermore, we recognise that the intervention we are testing resonate with the classical ‘information deficit model’. In its pure form, this model has received justifiable criticism for oversimplifying behaviour change - often in the context of paternalistic and culturally insensitive information provision.^36^ In contrast, our intervention is grounded in the lived experiences of those who have been unable to access care for want of basic information. By offering information at four points in time, including on the day of the appointment, we hoped to overcome the so-called ‘setting transfer problem’ that occurs when a participant has to remember their intention in a future time and space.^32,37^

### Primary outcome

Our primary outcome was the proportion of 18–44-year-olds attending a triage clinic either before, on, or up to 14 days after their appointed date (as previous programme data showed that some people miss their original appointment but attend to receive care in the following 2 weeks). A focus on left-behind groups is important to programme managers who are trying to close equity gaps, extend health service coverage, and ensure that their services do not exacerbate existing inequalities.

Our primary outcome was routinely collected by programme staff who check-in (register) every participant who presents to a treatment outreach clinic to receive care. Attendance was recorded using the Peek app, which automatically updates a central database that holds records of each participant’s eye care need, sociodemographic characteristics, arm allocation, and attendance status at the relevant treatment clinic on their appointed date. We used Bayesian methods to analyse the attendance data for every referred participant every 7 days and calculated the probability of attendance within each arm. We chose 7 days based on previous modelling and our expectation that approximately 300 people would be referred from the left-behind group every week.

To allow for the fact that the primary outcome includes attendance up to two weeks after the designated appointment date, and to ensure we had at least one week’s worth of data before conducting the first analysis, the date of the first analysis was set to occur 14 days after the latest scheduled appointment date among all participants enrolled during the first week of the study.

### Secondary outcome

If the intervention bundle was found to increase attendance among younger adults, we also wanted to check whether there had been an impact on the overall mean attendance rate across the entire population. This was to hedge against a situation where we adopted an intervention that improved access for the left-behind group but led to a large overall fall in attendance across the entire programme. As such, our secondary outcome was the proportion of people attending triage clinic on their appointed date across the entire population (i.e. aged 18-99 years old), again measured using routinely collected check-in data. We used the same outcome; attendance before, on, or up to 14 days after their appointed date.

### Sample size and stopping rules

As this was an adaptive trial, will did not pre-specify a sample size or estimate effect sizes for the intervention arms. Instead, participants were continually recruited until sufficient data accrued to trigger one of the two stopping rules. Triallists have argued that this approach is more “efficient, informative and ethical” than traditional fixed-design trials as this approach optimises the use of resources and can minimise the number of participants allocated to ineffective or less effective arms.^38^ Based on simulations, we selected the following rules:

1. There is a >95% probability that one arm is best, i.e. the difference between the two arms is >0%.
2. There is a >95% probability that the difference between the arms is <1% i.e. negligible.

### Simulations

Given that the marginal costs and programme impact associated with the intervention were negligible, we were willing to accept a relatively high type 1 error rate. This trade-off was made to increase the chance of correctly identifying a marginally more effective arm with a relatively small sample size. Based on Monte Carlo trial simulations, we expected a 36.3% chance of adopting the intervention when there was no true effect difference between it and the control. If the intervention increased attendance by 1%, we expected 81.0% chance of adopting the intervention. If the intervention increases attendance by 2% or more, we anticipated at least 90.5% chance of correctly adopting it. Further information is provided in the adaptive platform trial master protocol.^22^

### Implementation

As the trial was pragmatic, the responsibility for recruiting screening participants lay exclusively with the local programme team. Programme implementers enrolled participants by seeking consent from all those who required referral for further assessment and care (approximately 20% of all those screened).

Code within the Peek Vision screening platform used computer-generated random numbers to generate the allocation sequence and assign all consented, referred participants with equal numbers of participants in each arm, using even-numbered block sizes between 4-12. At the point of referral, the app displayed a colour-coded notice to the screener, telling them to read either the intervention or control script in the patient’s preferred language of English or Swahili. Software within the platform was programmed to autogenerate and send the enhanced reminder SMS on the same day, the day before the appointment, and on the appointment day to all those assigned to the intervention arm. Control arm participants received the autogenerated usual care SMS reminder on the day of referral and the day before the appointment.

Once assigned by the algorithm, each participant’s online record was automatically updated to display which arm they had been allocated to. Out of necessity, screeners and participants were not masked to assignment. Outcome assessment was performed by programme staff responsible for checking-in participants at triage clinic. No steps were taken to mask these staff to participant allocation status. Human investigators could not influence randomisation or allocation and had no role in performing statistical analyses: this was all performed by the trial algorithm. There were no plans to promote participant retention and complete follow-up.

The trial algorithm assessed whether the stopping rules had been met every 7 days, starting 14 days after the last appointment data for people enrolled during the first week. Figure 3 illustrates the trial algorithm process.

**Figure 3:**
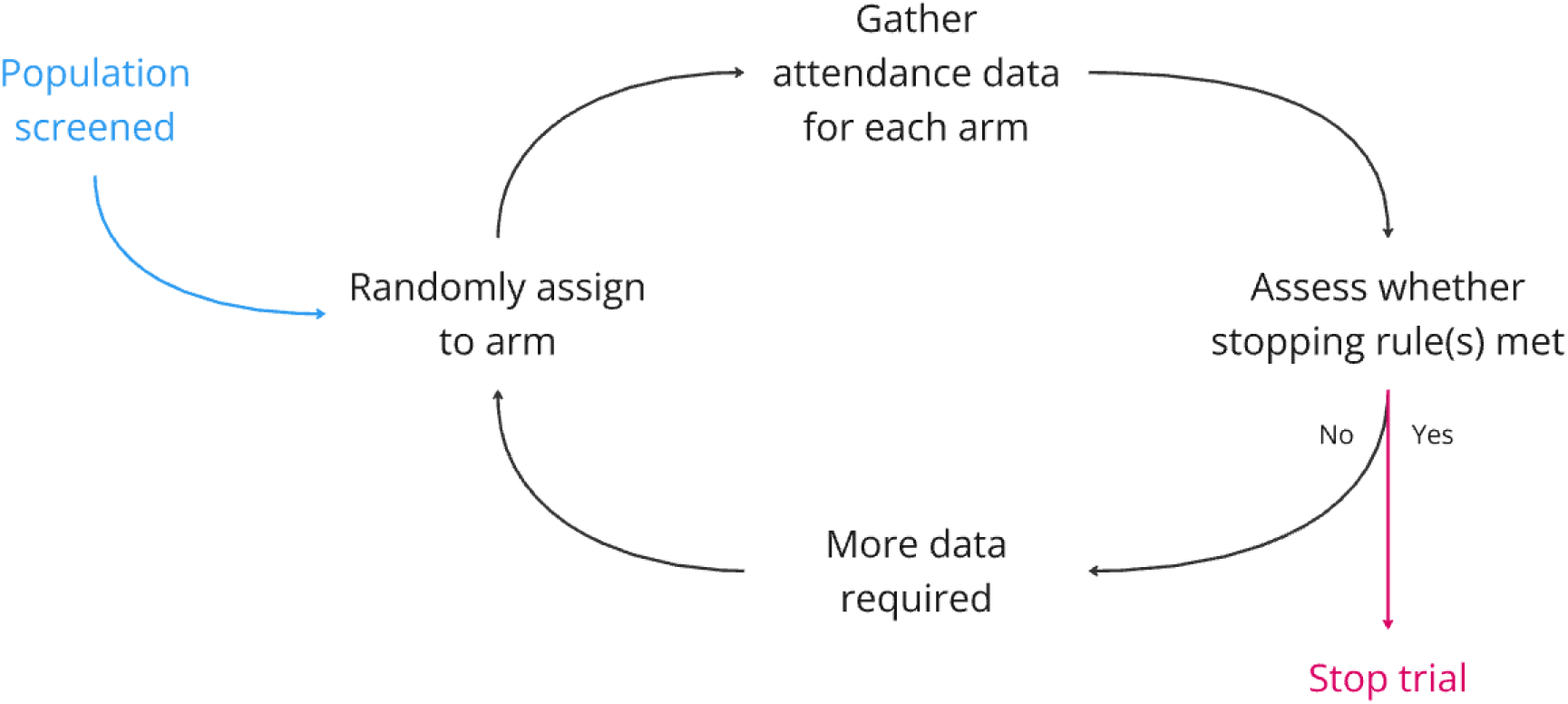
Algorithm process in executing the trial

### Statistical approach

Clinic attendance in each arm was assessed using Bayesian methods. At each analysis point, the total proportion of participants aged 18-44 (out of all accrued so far) attending in each arm was used to describe a binomial distribution of the outcome for each arm providing a distribution of the probability of attendance in each arm. This was combined with the prior distribution using Monte-Carlo simulations which generated the posterior probability distribution of the effect difference between the two arms. This posterior probability distribution was compared against the stopping rules, and if 95% of the posterior probability of the difference lay to one side of a difference of zero then the favoured arm would be declared superior and the trial stopped. If 95% of the posterior probability of the difference lay between +/-1% of zero then the arms would be declared equivalent and the trial stopped. A neutral prior was applied to provide the true state of data and allow the posterior distribution to be solely driven by the accumulating data.^39^

Upon completion of the trial, we performed our secondary analyses on an intention-to-treat basis. For the secondary outcomes, posterior probabilities were calculated for all participants, and again for participants aged >45 years, and for everyone who had been enrolled and randomised. We described the posterior distributions of the outcome proportions and the effect difference by reporting the means and their 95% credible intervals.

### Patient and public involvement

Lay people and a community advisory board reviewed and contributed to the development of this trial and the preceding work around identifying the left-behind group and identifying potential service improvements.

### Ethics

All participants provided informed written digital consent. The trial was approved by the KEMRI and LSHTM ethics committees.

## Findings

### Timeline and baseline characteristics

The trial started on 21^st^ May 2024. The superiority stopping rule was triggered at the first analysis point, ending enrollment to the trial on 20^th^ June 2024. By this date, a total of 2,143 younger adults had been screened and referred to local eye clinics, of whom 879 had consented and enrolled in the trial. Just over half (52.1%) of these enrolled 18-44-year-olds had been allocated to the control arm and 47.9% had been allocated to the intervention arm. Figure 4 illustrates the number of people randomized to each arm for the primary and secondary analyses.

**Figure 4:**
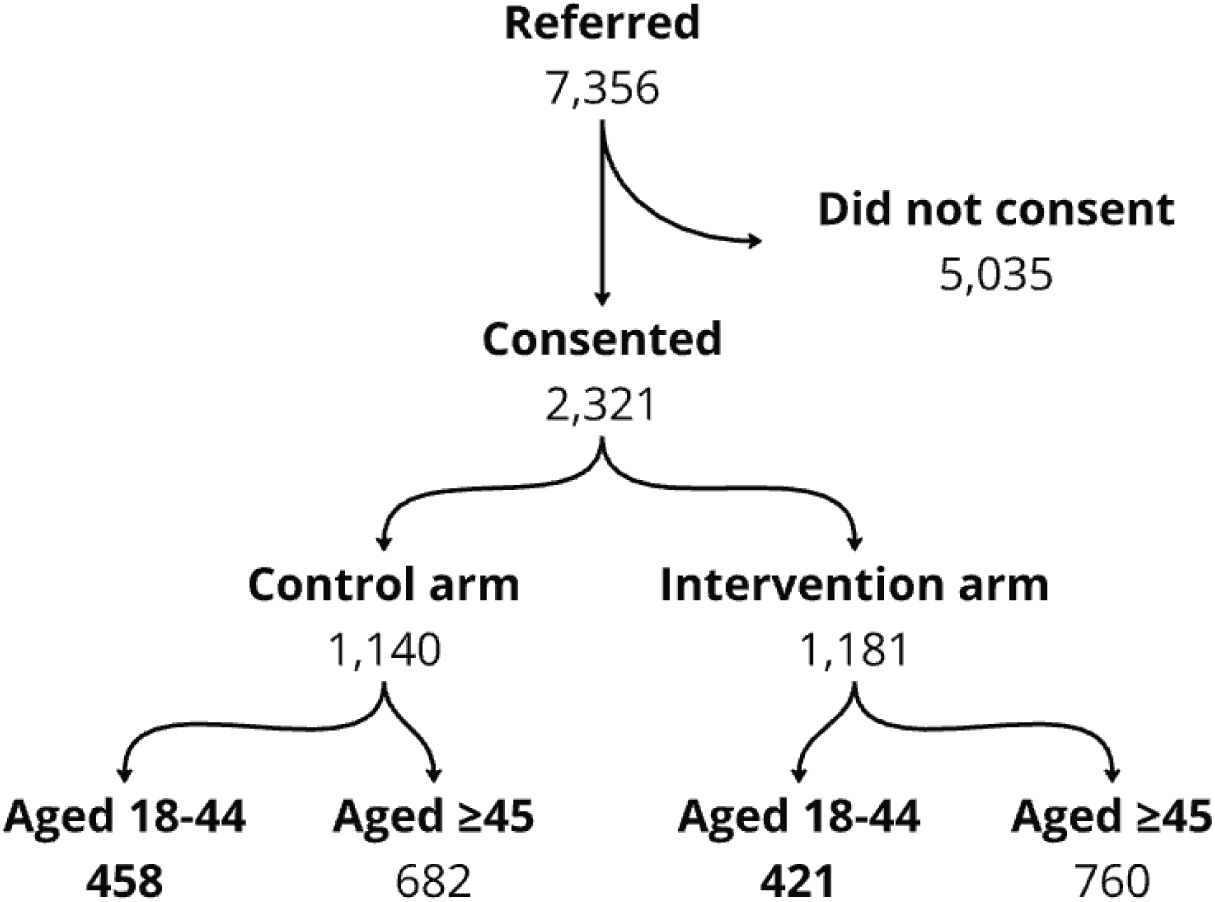
Randomisation to arms

Table 1 shows the baseline characteristics of the younger adults, older adults, and the entire population enrolled in the study. About two thirds of the consented participants were female. There were no losses to follow up or exclusions.

**Table 1:** Baseline characteristics

|  | 'Left behind' group (18-44 years) |  |  | Older adults (>45 years) |  |  | Entire population (18-99 years) |  |  |
| --- | --- | --- | --- | --- | --- | --- | --- | --- | --- |
|  | All | Intervention | Control | All | Intervention | Control | All | Intervention | Control |
| Number of participants | 879 | 421 | 458 | 1442 | 760 | 682 | 2321 | 1181 | 1140 |
| Mean age (SD) | 32.65 (7.87) | 32.76 (7.91) | 32.54 (7.85) | 60.56 (11.43) | 60.60 (11.28) | 60.52 (11.28) | 49.74 (17.20) | 50.48 (16.98) | 48.98 (17.39) |
| Female (%) | 624 (70.99) | 298 (70.78) | 326 (71.18) | 869 (60.26) | 473 (62.24) | 396 (58.06) | 1499 (64.17) | 774 (65.21) | 725 (63.10) |

### Primary analysis

At the first analysis point, 32.1% of younger adults in the control arm had accessed care vs 39.0% of younger adults in the intervention arm (Table 2). This resulted in an estimated 98.6% posterior probability that the intervention arm was superior, stopping the trial.

**Table 2:** Outcomes

|  | Total N | Accessed<br>care N<br>(%) | Probability<br>of being<br>superior | Probability of<br>having a<br>negligible<br>difference | Mean outcome<br>proportion<br>(95% CI)* | Mean effect<br>difference<br>(95% CI) |
| --- | --- | --- | --- | --- | --- | --- |
| 'Left behind' group: Younger adults (18-44 years) |  |  |  |  |  |  |
| Control arm | 458 | 147<br>(32.10%) | 1.38% | 2.96% | 32.14%<br>(27.91 - 36.46) | 6.37%<br>(0.59; 13.1) |
| Intervention arm | 421 | 164<br>(39.00%) | 98.62% |  | 38.98%<br>(34.39 - 43.70) |  |
| All adults (18-99 years) |  |  |  |  |  |  |
| Control arm | 1140 | 531<br>(46.58%) | 9.16% | 15.96% | 46.65<br>(43.64;49.39) | 2.77%<br>(-1.38;6.89) |
| Intervention arm | 1181 | 583<br>(49.36%) | 90.84% |  | 49.38%<br>(46.5;52.21) |  |

\* Note that the mean outcome proportion is based on a posterior distribution of outcome proportion and is therefore not exactly equal to the crude proportions because the prior moves the estimated proportion, ever so slightly, back towards the centre of the prior

### Secondary analysis

In the secondary analysis, conducted once the trial had stopped, we examined access rates across the entire population. We observed a 90.8% posterior probability that the intervention arm was superior. We note that the access rate in the control arm (46.6%) aligned with the pre-trial baseline access rate.

Based on concerns that the rise in attendance across the entire population randomised to the intervention arm might be purely driven by the rise in attendance among younger adults, we conducted a post-hoc analysis focused on outcomes in people aged exclusively >45 years. We observed a 27.51% probability that the difference between the intervention arm and the control arm was negligible (Table 3). Access rates were marginally lower in the intervention arm (55.1% vs 56.3% in the control arm).

**Table 3:** Outcomes among older adults (≥45 years)

|  | Total N | Accessed care N (%) | Probability of being superior | Probability of having a negligible difference | Mean outcome proportion (95% CI) * | Mean effect difference (95% CI) |
| --- | --- | --- | --- | --- | --- | --- |
| Control arm | 682 | 384 (56.3%) | 66.87% | 27.51% | 56.28% (52.50;60.04) | -1.15% (-6.33;4.04) |
| Intervention arm | 760 | 419 (55.1%) | 33.13% |  | 55.13% (51.62; 58.71) |  |

### Outcomes among those who attended on or before their due date

As our trial design used an outcome that allowed for people to attend their appointment up to 14 days late, our trial enrolled 4,164 participants, but only analysed data from 2,321 of these people. The remaining 1,843 had been enrolled, referred, randomised to an arm, and had the opportunity to attend their appointment early or on their due date, but were not included in the analysis because a full 14-day observation period had not yet elapsed since their appointment due date.

We noted that 94.7% of people in the primary analysis attended before/on their due date. We were able to use these additional data to triangulate our primary findings, using attendance on or before the due date as the outcome measure. We re-ran our analyses for younger adults, older adults, and the entire population, including everyone who had been randomised and whose appointment date fell on or before the date of the first analysis (20^th^ June 2024).

This post-hoc robustness check reinforced our primary findings. We found that the intervention had a 99.8% posterior probability of being superior among younger adults. The intervention had a 99.7% posterior probability of being superior among all adults, and a 70.1% posterior probability of being superior among older adults. Full results are presented in the Appendix.

### Implementation fidelity

During a routine interrogation of the data once the trial had ended, we found two fidelity issues. First, the intervention had not been implemented as intended due to a human error uploading the Swahili version of the intervention SMS. Everyone in the intervention arm received the enhanced verbal counselling and an extra reminder message as intended, and the 47.7% of participants in the intervention arm who spoke English received the enhanced SMS reminder message. However, the 52.3% of participants who spoke Swahili in the intervention arm had accidentally been sent the usual care SMS wording.

We conducted a post-hoc analysis to assess the effectiveness of the intervention on attendance by language, using the original primary outcome. We found that the intervention arm outperformed standard care in both language groups. Among Swahili speakers (i.e. those who received enhanced verbal counselling + one additional SMS reminder), there was a 98.8% probability that the intervention arm was superior. The probability was 80.6% among English speakers. Full results are presented in the Appendix.

Secondly, a bug in the code meant that a small proportion of the SMS messages due to be sent on the day of the appointment had been suppressed. This issue affected 32 people in the intervention arm. We performed a second sensitivity analysis that removed these people from the analysis. The outcomes were almost identical to the main intention-to-treat findings. Full results are presented in the Appendix.

## Discussion

In this embedded, pragmatic, adaptive RCT we found evidence that an intervention package suggested by members of the sociodemographic group facing the worst rates of access to essential eye services improved access for this group. We estimated that there is a 98.6% probability that enhanced counselling plus an extra reminder text message is superior to standard counselling and the standard number of reminder text messages among younger adults. We were able to adapt the clinical management software used by the local health system to autonomously execute several aspects of this adaptive trial, including randomising participants, delivering the SMS element of the intervention package, and analysing outcome data. The programme is now set up to conduct similar trials using the same approach.

Our finding that the interventions suggested by intended service beneficiaries was effective reinforces our belief that service improvement exercises should be grounded in the lived experience and perspectives of those the service is designed to serve. In the words of Turk et al: health service interventions ‘must be done with, and not simply done to, the people affected’.^31^ Community engagement is a core element of Primary Health Care,^40,41^ defined as ‘the process of involving people and communities in the design, planning and delivery of health services, thereby enabling them to make choices about care and treatment options or to participate in strategic decision-making on how health resources should be spent’.^42^ Our aspiration is that rapid qualitative engagement^12,13^ followed-up with rigorous embedded trials,^21^ become the engine of people-centred learning health systems.^43^ By focusing on left behind groups, our approach specifically aims to increase equitable access to care. WHO argue that ‘equity of access is central to UHC’.^44^

This study had a number of strengths and used an innovative study design. We used digital consenting, automated randomisation and allocation, autogenerated intervention delivery, automated statistical testing, an adaptive approach that used stopping rules to optimise sample size, routinely collected data, with the trial code embedded into the service delivery software platform. Lay representatives and wider stakeholders helped to design the study and interpret the findings, and the entire trial was conducted as part of a broader adaptive platform trial that enables rapid testing of new interventions as and when they emerge from ongoing qualitative engagement with left behind groups. Our post-hoc robustness checks delivered consistent findings.

Our study design also has a number of limitations. We had a relatively low consent rate, with more men declining to participate than women. While we can be confident that the intervention arm is superior, we need to bear in mind that there is a chance that the magnitude of difference between the arms is an overestimate because, on average, stopping rules end the trial at a local peak.^45^ This can be due to random deviations overestimating access to care under the intervention; underestimating access to care under standard care; or both. Encouragingly, we note that our previous equity analysis found a baseline attendance rate of 32% among 18-44-year-olds,^11^ which is almost exactly the same as the proportion observed in the control group, suggesting that it is unlikely that we have underestimated access to care in the control arm.

The posterior probability estimate calculated by our algorithm is sensitive to the choice of prior. Had we chosen a more sceptical prior, the probability estimates of superiority would have been lower for any given sample size. As such, our current estimates might overstate our true belief in the intervention’s effectiveness. However, as described in the methods, the stopping rule was set high (at 95%) to account for the uninformative prior, and we were comfortable with the level of type I error that our chosen methods were susceptible to. The underlying platform trial has been designed to optimise programmes by rapidly testing service modifications that are broadly equivalent to usual care in terms of costs, risks, and implementation feasibility. Under these conditions it is more important to avoid type II error than type I error, and we built in a relatively high acceptance rate for the latter. If we come to test interventions with meaningful cost or risk differentials in the future under this adaptive platform trial we would want to tighten the type I error rate.

SMS reminders have obvious limitations in the context of services for those with poor vision, as well as those living in rural setting without a reliable mobile phone networks or electricity. In addition, many people in Meru do not have a phone of their own. In the current programme, every screening participant who does not own a phone provides a contact number, and it may be that they can have the message read out to them. Additionally, all screening participants are given a referral card containing the appointment date, condition identified and the clinic’s name, complementing the SMS reminders. As our arms appear to have been well-balanced, we can assume that inability to receive or read an SMS message affected those in the intervention and control arms equally.

We chose to use a prioritarian approach that focuses on left-behind population groups. This prevented a situation where we would accept an intervention that improves the overall mean but was associated with a decline in access among left-behind groups, however this approach does not hedge against the slope of inequality worsening. Unfortunately, using a proportionate approach where we assess whether gains in each group are proportionate to their initial need would risk attributing success to our intervention rather than the more likely detection of regression toward the mean.

We used attendance as a proxy for access. Whilst this is the closest hard indicator available, the semantic implication of the term places responsibility on people rather than clinical systems or societal structures. We also note that we focus on a proximal indicator that does not always correlate well with receipt of high-quality care, or good clinical outcomes. We decided to focus on access for three main reasons; first it aligns with the conceptual narrative of Universal Health Coverage and ‘leaving no one behind’, second attendance data are already routinely collected and available for every single person who is referred, and third, internal Peek data suggests that the ‘fall off’ gap between those who are referred but do not attend is much larger than other gaps in the treatment cascade e.g. the proportion of those who attend but do not receive appropriate care, or the proportion of those who receive appropriate care but do not experience improved health outcomes.

Finally, a protocol violation meant that not everyone in the intervention arm received the full package of interventions. As such, whilst we can be confident that the combination of enhanced verbal counselling and sending an additional reminder on the day of the appointment is associated with higher attendance among younger adults, we do not know whether the enhanced SMS wording is effective as an individual component. Our study was not set up to answer this specific question. Finally, the intervention developed by young adults works well among this group but does not seem to make a meaningful impact on access rates for older adults.

## Conclusions

We deployed an adaptive trial that was embedded within an ongoing technology enabled screening programme. We used the trial architecture to test an intervention suggested by the sociodemographic group of intended service beneficiaries found to experience the worst rates of access. The enhanced counselling intervention package they suggested was found to boost access among this group. Future research should explore whether these innovative methods can be used to equitably improve access to other essential services and expand UHC.

## Data Availability

All data produced in the present study are available upon reasonable request to the authors.

## Disclaimer

The views expressed are those of the authors and not necessarily those of Wellcome, the NIHR or the Department of Health and Social Care.

## Contributors

LNA and MG are the study co-PIs. AB is the chief investigator. All authors jointly conceived the study, and analyses. DM and MK led the data analyses. All authors contributed to interpretation of the findings. LNA drafted the manuscript. All authors revised the manuscript and had final responsibility for the decision to submit for publication.

## Appendix

### 1. Post-hoc robustness check

Inclusion of all participants randomised to an arm, using attendance before or on appointment due data as the outcome.

#### Young adults 18-44 years

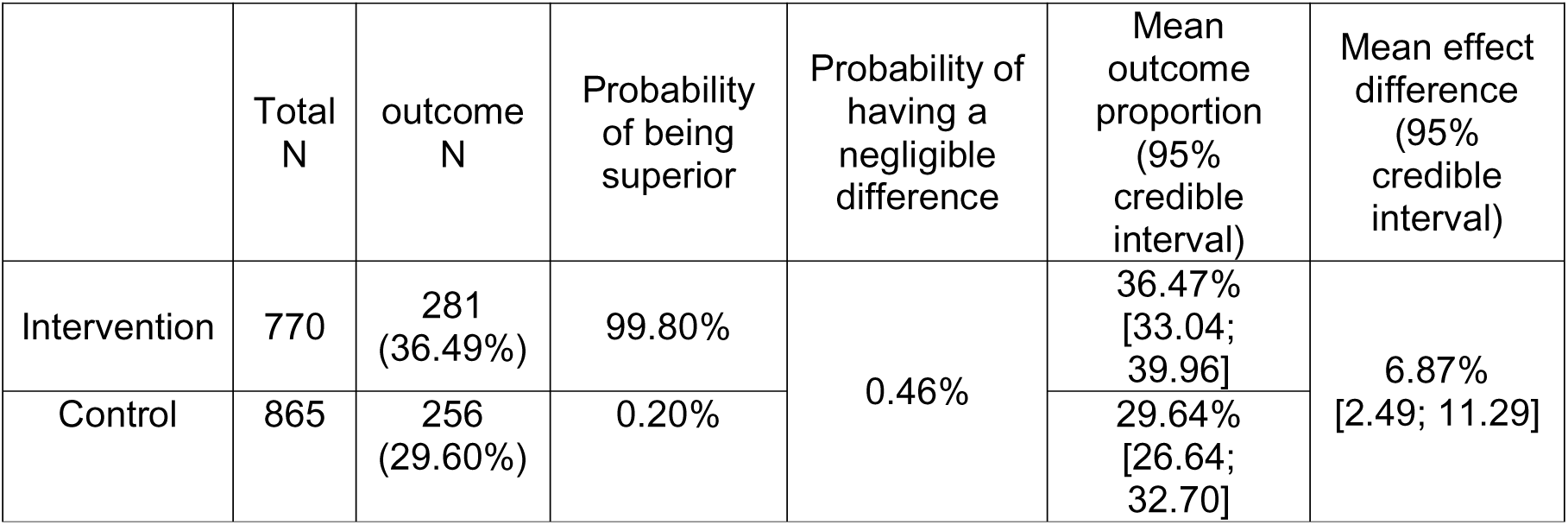

#### Older adults +44

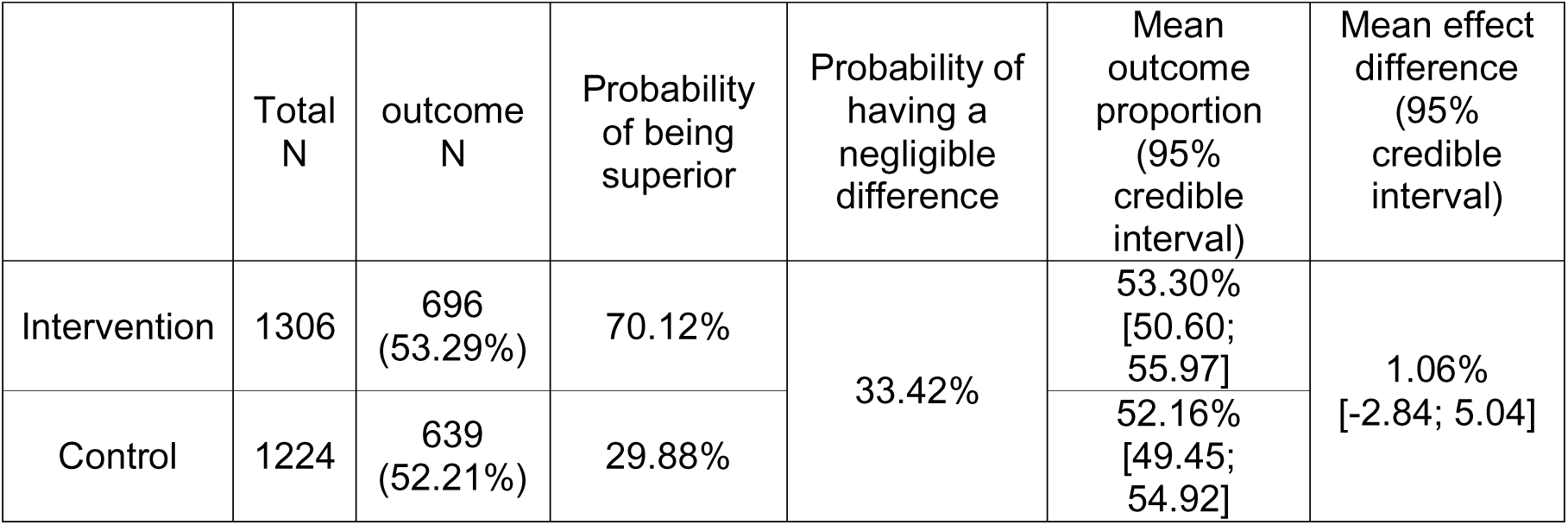

#### All adults

|  | Total N | outcome N | Probability of being superior | Probability of having a negligible difference | Mean outcome proportion (95% credible interval) | Mean effect difference (95% credible interval) |
| --- | --- | --- | --- | --- | --- | --- |
| Intervention | 2076 | 977 (47.06%) | 99.68% | 1.92% | 47.07% [44.91; 49.22] | 4.17% [1.13; 7.25] |
| Control | 2088 | 895 (42.84%) | 0.32% |  | 42.84% [40.77; 44.95] |  |

### 2. Effectiveness of the intervention on the primary outcome by language type

|  | Adults 18-44<br>English speakers |  |  |  | Adults 18-44<br>Swahili speakers |  |  |  |
| --- | --- | --- | --- | --- | --- | --- | --- | --- |
|  | Control |  | Intervention |  | Control |  | Intervention |  |
|  | 234/459<br>(51.0%) |  | 225/459<br>(49.0%) |  | 224/420<br>(53.3%) |  | 196/420<br>(46.7%) |  |
|  | Outcome | No outcome | Outcome | No outcome | Outcome | No outcome | Outcome | No outcome |
|  | 72/234<br>(30.7%) | 162/234<br>(69.2%) | 78/225<br>(34.7%) | 147/225<br>(65.3%) | 75/224<br>(33.5%) | 149/224<br>(66.5%) | 86/196<br>(43.9%) | 110/196<br>(56.1%) |
| Stop for efficacy? | No |  |  |  | Yes |  |  |  |
| Stop for futility? | No |  |  |  | No |  |  |  |

Among English speaking young adults, there was 80.6% posterior probability that the intervention arm was superior to the control.

Among Swahili speaking young adults, there was 98.8% posterior probability that the intervention arm was superior to the control.

At the point at which the trial was stopped, the intervention arm was outperforming the control arm among both language groups, but had only crossed the 95% threshold among Swahili speakers.

### 3. Effectiveness of the intervention on the primary outcome by trial fidelity

#### 1. Young adults 18-44 years

|  | Total N | outcome N | Probability of being superior | Probability of having a negligible difference | Mean outcome proportion (95% credible interval) | Mean effect difference (95% credible interval) |
| --- | --- | --- | --- | --- | --- | --- |
| Intervention | 412 | 160 | 98.19% | 3.06% | 38.81%<br>[34.18; 43.56] | 6.72%<br>[0.51; 13.2] |
| Control | 458 | 147 | 1.81% |  | 32.14%<br>[27.91; 36.46] |  |

#### 2. Older adults +44

|  | Total N | outcome N | Probability of being superior | Probability of having a negligible difference | Mean outcome proportion (95% credible interval) | Mean effect difference (95% credible interval) |
| --- | --- | --- | --- | --- | --- | --- |
| Intervention | 737 | 407 | 34.12% | 26.59% | 55.24%<br>[51.64; 58.79] | -1.12%<br>[-6.44; 4.15] |
| Control | 682 | 384 | 65.88% |  | 56.28%<br>[52.50; 60.04] |  |

#### 3. All adults

|  | Total N | outcome N | Probability of being superior | Probability of having a negligible difference | Mean outcome proportion (95% credible interval) | Mean effect difference (95% credible interval) |
| --- | --- | --- | --- | --- | --- | --- |
| Intervention | 1149 | 567 | 90.06% | 16.35% | 49.35%<br>[46.43; 52.25] | 2.73%<br>[-1.48; 6.97] |
| Control | 1140 | 531 | 9.94% |  | 46.65%<br>[43.64; 49.39] |  |

### 4. CONSORT checklist with relevant extensions

| Section | Item | Standard CONSORT Item | Equity extension | Pragmatic extension | Adaptive extension | Page |
| --- | --- | --- | --- | --- | --- | --- |
| **Title** |  |  |  |  |  |  |
| **Title** | 1a | Identification as a randomised trial in the title | If health equity is a major focus, consider using the term “health equity” in the title. |  |  | 1 |
| **Abstract** |  |  |  |  |  |  |
| **Structured Summary** | 1b | Structured summary of trial design, methods, results, and conclusions (for specific guidance see CONSORT for abstracts) | State research question(s) related to health equity |  |  | 2 |
|  | 1c |  | Present results of all planned health equity analyses |  |  | 2 |
|  | 1d |  | Describe extent and limits of applicability to populations of interest across PROGRESS-Plus characteristics |  |  | N/A |
| **Introduction** |  |  |  |  |  |  |
| **Background** | 2a | Scientific background and explanation of rationale | Describe rationale for focus on health equity | Describe the health or health service problem that the intervention is intended to address and other interventions that may commonly be aimed at this problem |  | 5-6 |
| **Objective** | 2b | Specific objectives or hypotheses | State the-objective being addressed with reference to health equity |  |  | 6 |
| **Methods** |  |  |  |  |  |  |
| **Trial Design** | 3a | Description of trial design (such as parallel, factorial) including allocation ratio | Describe aspects of trial design that were chosen to answer equity questions |  |  | 6 |
|  | 3b | Important changes to methods after trial commencement (such as eligibility criteria), with reasons |  |  | Important changes to the design or methods after trial commencement (such as eligibility criteria) outside the scope of the pre-planned adaptive design features, with reasons<br>Type of adaptive design used, with details of the pre-planned trial adaptations and the statistical information informing the adaptations | N/A<br>11 |
| **Participants** | 4a | Eligibility criteria for participants | Report population eligibility criteria across relevant PROGRESS-Plus | Eligibility criteria should be explicitly framed to show the degree to which |  | 7 |
|  |  |  | characteristics. | they include typical participants and/or, where applicable, typical providers (eg, nurses), institutions (eg, hospitals), communities (or localities eg, towns) and settings of care (eg, different healthcare financing systems) |  |  |
|  | 4b | Settings and locations where the data were collected | Report context and relationship to health inequity |  |  | 7 |
|  | 4c |  | Report details of partnerships with populations and communities, where applicable. |  |  | 7, 13 |
| **Intervention** | 5 | The interventions for each group with sufficient details to allow replication, including how and when they were actually administered | Report whether comparator intervention is the standard of care, and whether it has equity implications. | Describe extra resources added to (or resources removed from) usual settings in order to implement intervention. Indicate if efforts were made to standardise the intervention or if the intervention and its delivery were allowed to vary between participants, practitioners, or study sites<br>Describe the comparator in similar detail to the intervention |  | 7-8 |
| **Outcomes** | 6a | Completely defined pre-specified primary and secondary outcome measures, including how and when they were assessed | Report whether outcomes were identified as relevant and important to population(s) across PROGRESS-Plus characteristics and how this was done | Explain why the chosen outcomes and, when relevant, the length of follow-up are considered important to those who will use the results of the trial | Any other outcome measures used to inform pre-planned adaptations should be described with the rationale | 10 |
|  | 6b | Any changes to trial outcomes after the trial commenced, with reasons |  |  |  | 12? |
| **Sample Size** | 7a | How sample size was determined | Report whether analyses focused on health equity objectives are powered to detect differences. | If calculated using the smallest difference considered important by the target decision maker audience (the minimally important difference) then report where this difference was obtained | How sample size and operating characteristics were determined | 11 |
|  | 7b | When applicable, explanation of any interim analyses and stopping guidelines |  |  | Pre-planned interim decision-making criteria to guide the trial adaptation process; | 11 |
|  |  |  |  |  | whether decision-making criteria were binding or non-binding; pre-planned and actual timing and frequency of interim data looks to inform trial adaptations |  |
| **Randomisation Sequence Generation** | 8a | Method used to generate the random allocation sequence |  |  |  | 12 |
|  | 8b | Type of randomisation; details of any restriction (such as blocking and block size) | Report whether randomisation was stratified on PROGRESS-Plus characteristic(s) |  | Type of randomisation; details of any restriction (such as blocking and block size); any changes to the allocation rule after trial adaptation decisions; any pre-planned allocation rule or algorithm to update randomisation with timing and frequency of updates | 12 |
| **Allocation Concealment Mechanism** | 9 | Mechanism used to implement the random allocation sequence (such as sequentially numbered containers), describing any steps taken to conceal the sequence until interventions were assigned |  |  |  | 12 |
| **Implementation** | 10 | Who generated the random allocation sequence, who enrolled participants, and who assigned participants to interventions |  |  |  | 12 |
| **Blinding** | 11a | If done, who was blinded after assignment to interventions (for example, participants, care providers, those assessing outcomes) and how |  | If blinding was not done, or was not possible, explain why |  | 12 |
|  | 11b | If relevant, description of the similarity of interventions |  |  |  | 12 |
|  | 11c | [only applies for ACE] |  |  | Measures to safeguard the confidentiality of interim information and minimise potential operational bias during the trial | 12 |
| **Statistical Methods** | 12a | Statistical methods used to compare groups for primary and secondary outcomes |  |  | Statistical methods used to compare groups for primary and secondary outcomes, and any other outcomes used to make pre-planned adaptations | 12 |
|  | 12b | Methods for additional analyses, such as subgroup analyses and adjusted analyses | Report details of additional analyses focused on health equity, including whether analyses to estimate heterogeneity of effects between population subgroups were done on an additive or multiplicative scale, and whether pre-specified. |  |  | 10-12 |
|  | 12c | [ACE only] |  |  | For the implemented adaptive design features, statistical methods used to estimate treatment effects for key endpoints and to make inferences | 12 |
| **Ethical Concerns** | a | [equity only] | Report details of ethical clearance and informed consent |  |  |  |
| **Results** |  |  |  |  |  |  |
| **Participant flow (a diagram is strongly recommended)** | 13a | For each group, the numbers of participants who were randomly assigned, received intended treatment, and were analyzed for the primary outcome | Describe for each group, numbers of participants who were assigned, received and who were analyzed across relevant PROGRESS-Plus characteristics | The number of participants or units approached to take part in the trial, the number which were eligible, and reasons for non-participation should be reported | For each group, the numbers of participants who were randomly assigned, received intended treatment, and were analysed for the primary outcome and any other outcomes used to inform pre-planned adaptations, if applicable | 13 |
|  | 13b | For each group, losses and | Describe for each group, |  |  | 13 |
|  |  | exclusions after randomisation, together with reasons | losses and exclusions after randomisation across relevant PROGRESS-Plus characteristics, with reasons. |  |  |  |
| **Recruitment** | 14a | Dates defining the periods of recruitment and follow-up | Report whether methods of recruitment were designed to reach populations across relevant PROGRESS-Plus characteristics. |  | Dates defining the periods of recruitment and follow-up, for each group | 13 |
|  | 14b | Why the trial ended or was stopped |  |  |  | 13 |
|  | 14c | [ACE only] |  |  | Specify what trial adaptation decisions were made in light of the pre-planned decision-making criteria and observed accrued data | 13 |
| **Baseline Data** | 15 | A table showing baseline demographic and clinical characteristics for each group | Present the baseline characteristics also across relevant PROGRESS-Plus characteristics. |  |  |  |
|  | 15b | [ACE only] |  |  | Summary of data to enable the assessment of similarity in the trial population between interim stages |  |
| **Numbers Analyzed** | 16 | For each group, number of participants (denominator) included in each analysis and whether the analysis was by original assigned groups |  |  |  |  |
| **Outcomes and Estimation** | 17a | For each primary and secondary outcome, results for each group, and the estimated effect size and its precision (such as 95% confidence interval) |  |  |  |  |
|  | 17b | For binary outcomes, presentation of both absolute and relative effect sizes is recommended |  |  |  |  |
|  | 17c | [ACE only] |  |  | Report interim results used to inform interim decision-making |  |
| **Ancillary Analysis** | 18a | Results of any other analyses performed, including subgroup analyses and adjusted analyses, distinguishing pre-specified from exploratory | Give the results of additional analytic approaches related to equity objectives distinguishing pre-specified from exploratory. |  |  |  |
|  | 18b |  | Details of implementation (coverage, intensity) in each trial arm across relevant PROGRESS-Plus characteristics |  |  |  |
| **Harms** | 19 | All important harms or unintended effects in each group (for specific guidance see CONSORT for harms ) | Report whether intervention generated inequities (e.g. unintended effects) were assessed |  |  |  |
| **Discussion** |  |  |  |  |  |  |
| **Limitation** | 20 | Trial limitations, addressing sources of potential bias, imprecision, and, if relevant, multiplicity of analyses | Report any limitations related to assessing effects on health equity. |  |  |  |
| **Generalizability** | 21 | Generalisability (external validity, applicability) of the trial findings | In addition, report applicability related to population of interest across PROGRESS-Plus characteristics. | Describe key aspects of the setting which determined the trial results. Discuss possible differences in other settings where clinical traditions, health service organisation, staffing, or resources may vary from those of the trial |  |  |
| **Interpretation** | 22 | Interpretation consistent with results, balancing benefits and harms, and considering other relevant evidence |  |  |  |  |
| **Other Information** |  |  |  |  |  |  |
| **Registration** | 23 | Registration number and name of trial registry |  |  |  |  |
| **Protocol** | 24 | Where the full trial protocol can be accessed, if available |  |  |  |  |
|  | 24b | [ACE only] |  |  | Where the full statistical analysis plan and other relevant trial documents can be accessed |  |
| **Funding** | 25 | Sources of funding and other support (such as supply of drugs), role of funders |  |  |  |  |

ACE, Adaptive designs CONSORT Extension

